# Meta-Analyses of Sexual Assault Prevalence Among Homeless Women

**DOI:** 10.64898/2026.03.14.26348410

**Authors:** Sarah Jean Valliant, Jorah Razumeyko, Andreia Silva, Samantha S. Parton, Angelina Ka Lee, Jacob Derin, Noor Banihashem Ahmad, Carole M. Kulik, Mary Banihashem

## Abstract

**Background:** Literature on sexual assault prevalence among homeless women is limited, with few studies disaggregating risk by geography, resource access, mental health, LGBTQ status, or disability.

**Objective:** This study provides two distinct meta-analyses to ascertain the aggregated overall prevalence (k=20 studies) and the aggregated 12-month prevalence (k=14 studies) of sexual assault among homeless women. By examining each recall period independently, we elucidate cumulative burden throughout the life cycle and annual risk, offering unique insights for public health interventions. By synthesizing global data, we aimed to clarify risks for women with disabilities, mental illness, or Lesbian, Gay, Bisexual, Transgender, Queer or Questioning, Plus (LGBTQ+) identities to inform crisis care interventions.

**Methods:** Following PRISMA 2020 guidelines, six databases were searched for studies published after 2010 reporting sexual assault prevalence in homeless women. Twenty studies met the inclusion criteria. Random-effects meta-analyses were performed using a logit transformation. Heterogeneity was assessed with I² and Cochran’s Q; publication bias with funnel plots and Egger’s test.

**Results:** The pooled lifetime prevalence of sexual assault was 39.2 % (95 % CI 25–56 %), and 12-month prevalence was 22 % (95 % CI 16–30 %). Heterogeneity was extreme (I² = 97 %). Subgroup analyses showed the highest prevalence among women with disabilities (92 %, single study), followed by LGBTQ+ (33 %) and women with mental illness (34 %). HIV-positive women had the lowest prevalence (2.6 %). Egger’s test indicated no publication bias (p = 0.64).

**Conclusion:** Homeless women face disproportionately high rates of sexual assault, far exceeding the general female population, with particularly elevated estimates among women with disabilities, LGBTQ+ women, and those with mental illness. These preliminary findings highlight the need for improved screening practices and tailored public health interventions to address sexual assault in doubly vulnerable populations. Standardizing definitions of sexual assault and investigating risk factors could lead to more tailored public health interventions.

**Highlights:**

- Marked Epidemiologic Burden: Nearly 40% of homeless women report lifetime sexual assault.
- Persistent risk: One in five homeless women assaulted within the past 12 months.
- Marginalized Impact: Rates highest among disabled, LGBTQ+, and HIV+ women.
- High Variability: Extreme heterogeneity (I² ≈ 97%) shows research inconsistency.
- Research Priority: Standardize methods and definitions to improve accuracy.

## 1. Introduction

### 1.1. Rationale

Sexual violence is a pervasive public health issue [1]. Globally, approximately one in four women experiences physical and/or sexual violence in their lifetime [2]. In the United States, data from the 2012 National Intimate Partner and Sexual Violence Survey indicates that approximately 19.3% of women report a lifetime history of sexual assault, with 12-month prevalence in the general population averaging 1.6% [2,3]. These statistics highlight that while sexual assault is not uncommon, it is typically less frequent in the general population on a year-to-year basis. By contrast, homeless women face multiple overlapping vulnerabilities, including financial instability, unsafe living environments, and limited access to social and medical resources that increase their risk of violence [4]. Prevalence estimates in this population vary widely.

Such variability suggests that sexual assault among homeless women is both more common and more heterogeneous than in the general population. Comparisons across studies were further complicated by differences in study design, geographic context, and sample composition [5]. Few studies disaggregate risk by disability status, mental illness, or LGBTQ+ identity [6], and definitions of sexual assault are often inconsistent across survey instruments [7]. Given this heterogeneity and the limited subgroup analysis available, a meta-analysis was needed to clarify the overall burden of sexual assault among homeless women and to inform targeted public health interventions. Pooled prevalence estimate offers policymakers and health agencies a clear understanding of the scope, with a clearer picture of sexual assault among homeless women, beyond the fragmented results of individual studies [8]. Synthesizing data across regions and methodologies also strengthens the evidence base, producing more reliable estimates to guide intervention and service design [9]. Importantly, meta-analysis allows identification of high-risk subgroups, such as women with disabilities, mental illness, or LGBTQ+ identities, which is critical for targeted prevention strategies and resource allocation [10].

Among the many forms of violence experienced by homeless women, sexual assault constitutes one of the most pervasive threats. Homeless women are victimized at higher rates than the average population of women [4]. Even within the homeless populations, unsheltered women face elevated risks. Studies suggest that 57% of unsheltered homeless women in Los Angeles reported being physically assaulted, 28% more than those in shelters. That robbery was reported by 73% of unsheltered versus 28% of sheltered women. These differences remained significant even after controlling for mental health, demographic factors, and substance use [11]. Sexual victimization has long-lasting consequences, including physical injury, unintended pregnancy, heightened risk of sexually transmitted infections, and enduring mental health conditions such as depression, anxiety, post-traumatic stress disorder, and substance misuse. Despite recognition of the heightened risks faced by homeless women, the intersectional risk factors for sexual assault have been difficult to ascertain, owing to methodological inconsistencies across studies, variability in definitions of sexual assault or homelessness, and a lack of interest in researching specific risk factors. Intersectional risk factors refer to the overlapping characteristics that may or may not increase vulnerability among homeless women; in this study, we focused on mental health conditions, sexuality and gender identity, HIV status, and physical disability [12].

A growing body of research in North America and internationally shows that women experiencing homelessness face disproportionately high rates of sexual violence, often far exceeding those in the general population [2]. Community-based surveys conducted in Los Angeles and San Francisco documented sexual victimization as a frequent experience among women utilizing shelters, street-based services, or community drop-in centers [3]. Studies of homeless youth similarly report high rates of survival sex and coerced encounters used to secure basic resources [8]. At the same time, research among rural and Indigenous populations highlights compounded vulnerability from structural marginalization, racism, and limited service access [2]. However, the literature remains fragmented.

Many studies employ small, nonprobability samples drawn from service-using populations [7], including individuals who receive assistance, such as shelter, food programs, or other formal support services [7]. This approach may potentially underestimate or overestimate prevalence rates compared to those not connected to formal support services [7, 9]. Recall periods, a method researchers use when participants self-report past experiences, vary substantially, with some studies examining lifetime exposure to sexual assault. In contrast, other studies focus on recent or twelve-month prevalence, which complicates direct comparisons.

Operationalization of sexual assault likewise differs, ranging from narrow legalistic definitions to broader measures encompassing coercion, attempted assault, or transactional sex [4]. Moreover, reporting is limited by stigma, mistrust of institutions, and fear of involvement in the criminal justice system. Women with disabilities usually face heightened vulnerability due to dependence on caregivers, barriers to reporting abuse, and pervasive discrimination.

LGBTQ+ women frequently encounter elevated risks related to rejection, transphobia, and survival sex, yet their experiences remain under-examined in many studies [8]. Similarly, women with severe mental illness constitute a subgroup subject to intersecting stigma and social isolation, which may exacerbate risk and undermine access to protective services [4]. Few studies have disaggregated findings to assess whether these populations face disproportionately higher prevalence of sexual assault, leaving a critical gap in knowledge that undermines equity-oriented service provision [3].

Within this context, a meta-analysis & synthesis of the available evidence is warranted. Meta-analytic techniques offer an opportunity to integrate heterogeneous studies, derive pooled prevalence estimates, and formally assess subgroup variation. By quantifying the burden of sexual assault among homeless women, such an analysis offers essential epidemiological grounding for prevention, policy, and clinical response. Recognition of subgroup disparities is pivotal for guiding the development of interventions that are responsive to heterogeneity in risk, thereby safeguarding against the marginalization of those with overlapping vulnerabilities in policy and program implementation.

The existing literature on sexual assault among homeless women is characterized by highly variable prevalence estimates, complicated by differences in study design, definition of sexual assault, geographic context, and recall period. Such variability has made it difficult to ascertain the overall public health burden. A meta-analysis is therefore warranted not only to derive a pooled prevalence estimate for policymakers, but more critically, to formally quantify and investigate the reasons for this extensive heterogeneity. In addition to performing a meta-analysis for a standardized recall period of 12 months, our approach also deliberately synthesizes studies with varied recall periods, ranging from 30 days to lifetime, to calculate a comprehensive overall prevalence that reflects the cumulative risk factors many homeless women experience. This study employs a random-effects model, subgroup analyses, and meta-regression to transform heterogeneity between studies into a key finding, rather than a methodological challenge. Exploring the systematic factors that influence risk & reporting across diverse settings could support future public health research and policy implementation. This meta-analysis directly contributes to Heliyon’s mission to synthesize multidisciplinary public-health evidence.

### 1.2 Objectives

The present study addresses inconsistent prevalence estimates through a systematic review and meta-analysis of 20 studies reporting on sexual assault among homeless women. The study pooled lifetime and 12-month prevalence of sexual assault among homeless women to determine which subgroups face the highest risk, to inform crisis response and tailored public health interventions. Specifically, the study aimed to:

1. Estimate the pooled prevalence of sexual assault across diverse study designs and settings.
2. Compare prevalence estimates by recall period, focusing on lifetime versus 12-month experiences where available;
3. Examine subgroup differences among women with disabilities, LGBTQ+ populations, and women with mental illness; and
4. Evaluate the extent of heterogeneity and potential publication bias within the available literature.

In doing so, this study seeks to provide a more robust and comprehensive understanding of the epidemiology of sexual assault in homeless women, thereby informing clinical practice, public health initiatives, and structural interventions to reduce violence against one of society’s most vulnerable populations.

## 2. Methods

### 2.1 Eligibility Criteria

Eligible studies focused on women experiencing homelessness, with homelessness explicitly defined (e.g., living on the streets, in shelters, or in temporary accommodations). Studies were required to report prevalence rates of sexual assault or sexual violence, with “sexual assault” defined as forced penetration (rape), unwanted sexual contact, or coercion through threats, intimidation, or manipulation. Acceptable study designs included peer-reviewed original research articles using observational designs (cross-sectional, cohort, or case–control) or interventional studies that provided baseline prevalence data. To ensure comparability, studies must give sufficient information for the extraction of prevalence rates or effect-size measures, including raw numbers, percentages, odds ratios, or relative risks. The certainty and robustness of both the lifetime and 12-month prevalences were evaluated through a systematic risk-of-bias assessment and multiple sensitivity analyses.

Exclusion criteria are predefined. Studies are excluded if they did not focus on women or disaggregate data for homeless women, did not provide precise prevalence estimates, or reported only risk factors or qualitative data without accompanying prevalence estimates. Anecdotal reports, commentaries, editorials, non-peer-reviewed literature, conference abstracts, and studies using retrospective data before 2010 are excluded. Studies that combined sexual harassment under the umbrella of sexual violence without clear definitions are also excluded.

### 2.2 Information Sources

A systematic search was conducted across six databases: PubMed, Directory of Open Access Journals, PsycInfo, OpenGrey, Bielfeld, OneSearch, and Google Scholar. The search included peer-reviewed studies published after 2010 to ensure inclusion of recent, relevant data. Additionally, only English articles are utilized in the analysis.

### 2.3 Search Strategy

A comprehensive literature search was conducted to identify peer-reviewed studies reporting the prevalence of sexual assault among women experiencing homelessness. Six electronic databases were systematically searched: PubMed, Directory of Open Access Journals (DOAJ), PsycInfo, OpenGrey, Bielefeld Academic Search Engine (BASE), OneSearch, and Google Scholar. The search was conducted from June 2025 to October 2025 and limited to English-language articles published from January 2010 onward, to ensure inclusion of recent, methodologically comparable research.

Search strings were constructed using controlled vocabulary (MeSH terms) and free-text keywords relevant to homelessness, gender, and sexual violence. The Boolean operators *AND* and *OR* were applied to combine key terms. An example search string used in PubMed was: “homeless women” OR “unhoused women” OR “women experiencing homelessness” AND “sexual assault” OR “sexual violence” OR “rape” OR “coerced sex”.

Reference lists of all included articles were also manually screened to identify additional eligible studies. Search procedures followed PRISMA 2020 guidelines to ensure transparency and reproducibility.

### 2.4 Study Selection

The study selection process occurred in two phases. In the first phase, three independent reviewers screened titles and abstracts for relevance using the inclusion and exclusion criteria. Potentially eligible studies underwent full-text review. In the second phase, full texts were reviewed, and relevant information was documented in a structured data-extraction form. A comprehensive search across seven electronic databases yielded 164 records. Following the removal of 46 duplicate entries, 118 unique studies remained and were advanced for title and abstract screening. Of these, 38 underwent full-text review, and 20 met the inclusion criteria for analysis. The study sample comprised participants from multiple countries, including 18 from the United States, one from Spain, and one from France, reflecting a predominantly U.S.-based distribution. Discrepancies were resolved through discussion and consensus with a supervising investigator.

### 2.5 Data Collection Process

Data was extracted independently by reviewers using standardized forms. Extracted information included study characteristics (author, year, country, design, methodology, sample size, and URLs), definitions of homelessness, and demographic details of participants. Populations were classified into four groups: unhoused women who experienced sexual assault, unhoused women who had not experienced sexual assault, housed women who experienced sexual assault, and housed women who had not experienced sexual assault. Additional comorbidities were recorded when available. Sexual assault outcomes were defined by this meta-analysis as self-reported or non-self-reported prevalence rates, with breakdowns by subgroup (e.g., LGBTQ+, pregnant women, women with disabilities). Sexual assault was categorized by type (e.g., physical assault, rape, coercion) and timeframe (lifetime, 12-month, or single incident).

Statistical and methodological data from the studies were also extracted, including effect sizes (odds ratios and risk ratios), confidence intervals, p-values, and the statistical tests used. Risk-of-bias assessments for each study are presented in Table 2. All included studies underwent independent quality assessment by at least three research assistants. Studies flagged for secondary review underwent a more detailed data extraction process to ensure accuracy and completeness. For these studies, reviewers recorded comprehensive study characteristics (eg, author, year, country, design, sample size, and URLs), along with population classifications, definitions of homelessness, demographic details, and comorbidities. Sexual assault outcomes were extracted with greater granularity, including subgroup prevalence, assault type, and timeframe. Methodological features and risk-of-bias assessments were re-evaluated, with discrepancies resolved by consensus under the supervision of a senior investigator.

### 2.6 Data Items

Table 1 summarizes the categories and variables extracted from the included studies. The data extraction process encompassed study characteristics, population demographics, sexual assault outcomes, statistical methods, and secondary review data to ensure methodological rigor and completeness.

**Table 1:** All data extraction types are present in Table 2.

| *Category* | *Data Extracted \** |
| --- | --- |
| Study Characteristics | Author(s), year of publication, study location, research design, methodology, sample size, and links/URLs to access the study. |
| Population Characteristics | Participants' demographics, definition of homelessness, and population classifications. |
| Sexual Assault Outcomes | Prevalence rates of sexual assault. |
| Statistical Methodological Data | Extracted effect sizes (OR, RR), CI, P-value, stats test used, and any reported risk-of-bias assessment. |
| Secondary Review Data | All studies are required for additional review and re-examination to ensure completeness and accuracy. |

\* Includes methodology for extractions, including what values were extracted.

**Table 2:**
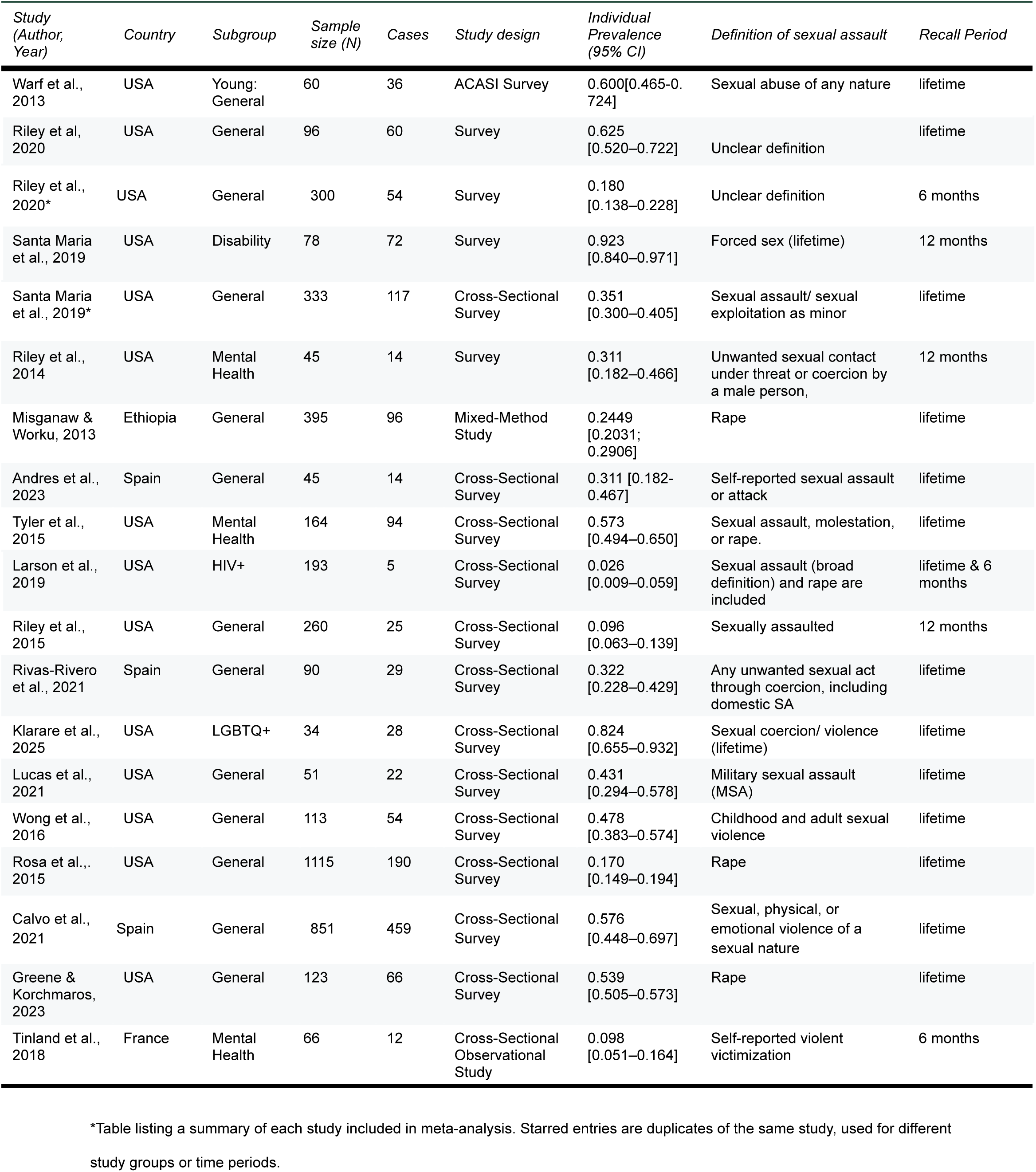
Titles of the 20 studies used to determine the proportion of sexual assault prevalence among homeless women.

| *Study (Author, Year)* | *Country* | *Subgroup* | *Sample size (N)* | *Cases* | *Study design* | *Individual Prevalence (95% CI)* | *Definition of sexual assault* | *Recall Period* |
| --- | --- | --- | --- | --- | --- | --- | --- | --- |
| Warf et al., 2013 | USA | Young: General | 60 | 36 | ACASI Survey | 0.600[0.465-0.724] | Sexual abuse of any nature | lifetime |
| Riley et al., 2020 | USA | General | 96 | 60 | Survey | 0.625 [0.520–0.722] | Unclear definition | lifetime |
| Riley et al., 2020* | USA | General | 300 | 54 | Survey | 0.180 [0.138–0.228] | Unclear definition | 6 months |
| Santa Maria et al., 2019 | USA | Disability | 78 | 72 | Survey | 0.923 [0.840–0.971] | Forced sex (lifetime) | 12 months |
| Santa Maria et al., 2019* | USA | General | 333 | 117 | Cross-Sectional Survey | 0.351 [0.300–0.405] | Sexual assault/ sexual exploitation as minor | lifetime |
| Riley et al., 2014 | USA | Mental Health | 45 | 14 | Survey | 0.311 [0.182–0.466] | Unwanted sexual contact under threat or coercion by a male person, | 12 months |
| Misganaw & Worku, 2013 | Ethiopia | General | 395 | 96 | Mixed-Method Study | 0.2449 [0.2031; 0.2906] | Rape | lifetime |
| Andres et al., 2023 | Spain | General | 45 | 14 | Cross-Sectional Survey | 0.311 [0.182-0.467] | Self-reported sexual assault or attack | lifetime |
| Tyler et al., 2015 | USA | Mental Health | 164 | 94 | Cross-Sectional Survey | 0.573 [0.494–0.650] | Sexual assault, molestation, or rape. | lifetime |
| Larson et al., 2019 | USA | HIV+ | 193 | 5 | Cross-Sectional Survey | 0.026 [0.009–0.059] | Sexual assault (broad definition) and rape are included | lifetime & 6 months |
| Riley et al., 2015 | USA | General | 260 | 25 | Cross-Sectional Survey | 0.096 [0.063–0.139] | Sexually assaulted | 12 months |
| Rivas-Rivero et al., 2021 | Spain | General | 90 | 29 | Cross-Sectional Survey | 0.322 [0.228–0.429] | Any unwanted sexual act through coercion, including domestic SA | lifetime |
| Klarare et al., 2025 | USA | LGBTQ+ | 34 | 28 | Cross-Sectional Survey | 0.824 [0.655–0.932] | Sexual coercion/ violence (lifetime) | lifetime |
| Lucas et al., 2021 | USA | General | 51 | 22 | Cross-Sectional Survey | 0.431 [0.294–0.578] | Military sexual assault (MSA) | lifetime |
| Wong et al., 2016 | USA | General | 113 | 54 | Cross-Sectional Survey | 0.478 [0.383–0.574] | Childhood and adult sexual violence | lifetime |
| Rosa et al., 2015 | USA | General | 1115 | 190 | Cross-Sectional Survey | 0.170 [0.149–0.194] | Rape | lifetime |
| Calvo et al., 2021 | Spain | General | 851 | 459 | Cross-Sectional Survey | 0.576 [0.448–0.697] | Sexual, physical, or emotional violence of a sexual nature | lifetime |
| Greene & Korchmaros, 2023 | USA | General | 123 | 66 | Cross-Sectional Survey | 0.539 [0.505–0.573] | Rape | lifetime |
| Tinland et al., 2018 | France | Mental Health | 66 | 12 | Cross-Sectional Observational Study | 0.098 [0.051–0.164] | Self-reported violent victimization | 6 months |

\*Table listing a summary of each study included in meta-analysis. Starred entries are duplicates of the same study, used for different study groups or time periods.

### 2.7 Study Risk of Bias Assessment

Risk of bias was evaluated using the Newcastle–Ottawa Scale, which was applied to all eligible studies. A minimum of three reviewers independently assessed each study. Discrepancies were resolved through consensus.

Two of the principal risks of bias across both studies are underrepresentation and recall bias in the self-reported Measures. Due to the reliance on convenience sampling, the sample size may underrepresent unsheltered women, thereby limiting the generalizability of the results. Additionally, using self-reported measures of sexual assault introduces the risk of reporting and recall bias. Overall, for the self-reporting assessment, the existence of inconsistent definitions of sexual violence, along with the variability of interpretation in measurement tools, can further complicate comparability.

Although the meta-analysis exhibits exceptionally high heterogeneity, the systematic risk-of-bias assessment concluded that the findings are robust. The results are not significantly skewed by publication bias or the undue influence of any single study, lending confidence to the overall conclusion of a high prevalence of sexual assault among homeless women. No significant evidence of publication bias was detected, indicating that the pooled results are likely representative of the available literature and strengthening the validity and reliability of the meta-analytic conclusions for public health interpretation.

### 2.8 Effect Measures

The primary effect measure was the pooled prevalence proportion of sexual assault among homeless women. For each study, prevalence (p) was defined as the number of homeless women who reported experiencing sexual assault divided by the total number of homeless women included in the study sample. To stabilize the variance of proportions, individual study estimates were transformed using a logit transformation before pooling. Pooled prevalence and corresponding 95% confidence intervals (CIs) were calculated under a random-effects model using the DerSimonian–Laird estimator to account for between-study heterogeneity. Heterogeneity was quantified using the Q statistic, I², and τ² values.

Publication bias was visually assessed using funnel plots and statistically tested through Egger’s regression test. Egger’s test results (p = 0.64 and 0.73) suggest no evidence of publication bias; however, caution is warranted as the test has limited statistical power when fewer than ten studies are included.

### 2.9 Synthesis Methods

Both qualitative and quantitative syntheses were executed. For the qualitative synthesis, a narrative approach was used to summarize and compare study characteristics, prevalence estimates, and subgroup-specific risks. Themes such as definitions of homelessness, methodological approaches, and reported risk factors are highlighted, with particular focus on differences across subgroups.

Two complementary quantitative syntheses were completed. The first, referred to as the “lifetime prevalence” synthesis, combined all 20 eligible studies to generate a broad estimate of overall risk, regardless of the recall period used (ranging from 30 days to lifetime). The second, more targeted analysis drew on a subset of 14 studies that specifically reported prevalence within a 12-month or shorter timeframe. This dual strategy was adopted to provide a more refined estimate of recent sexual assault prevalence and a detailed overview of the available evidence.

For quantitative synthesis, studies that reported sufficiently homogeneous data were included in a meta-analysis. Pooled prevalence estimates were calculated for overall sexual assault prevalence among homeless women. High-risk subgroups, including women with disabilities, LGBTQ+ women, and women with mental illness, had a greater influence on the pooled prevalence. Effect sizes were calculated directly; where necessary, odds ratios were converted to risk ratios to allow comparability. For studies reporting only raw numbers, prevalence and effect sizes were estimated.

Heterogeneity was assessed using Cochran’s Q and quantified with the I² statistic. Heterogeneity thresholds followed the Cochrane conventions: 25% = low, 50% = moderate, 75% = high. When heterogeneity was low, a fixed-effects model was used; when it was moderate to high, a random-effects model was applied. Subgroup and meta-regression analyses were conducted to identify sources of heterogeneity and disentangle subgroup effects. Publication bias was evaluated using funnel plots and Egger’s test. Sensitivity analyses were performed to test the robustness of the findings.

### 2.10 Reporting Bias Assessment

The potential for reporting bias and small-study effects was evaluated for both the lifetime and 12-month prevalence estimates using a combination of visual and statistical approaches. Funnel plots were visually inspected to detect asymmetry that might indicate publication bias. In addition, Egger’s linear regression test was performed to formally assess funnel plot asymmetry. To further evaluate the robustness of the pooled prevalence estimates, a sensitivity analysis using the trim-and-fill method was conducted, which estimates the number of potentially missing studies and adjusts the overall effect size accordingly. These complementary approaches provide confidence that the reported prevalence estimates are not unduly influenced by selective reporting or small-study effects.

### 2.11 Certainty Assessment

The certainty and robustness of both the lifetime and 12-month prevalence estimates were assessed through a systematic review of risk of bias using established criteria across key domains, including selection, reporting, and measurement bias. Heterogeneity among studies was quantified using I² and τ² statistics, and multiple sensitivity analyses were conducted to evaluate the stability of the pooled estimates. Leave-one-out analyses tested whether the overall pooled estimate was disproportionately influenced by any single study, while Baujat plots visually identified the studies that contributed most to heterogeneity. Additional influence diagnostics, including studentized residuals, DFFITS, Cook’s distance, leverage, and τ² deletion, were used to detect studies or subgroups exerting undue influence.

Studies flagged by these analyses were further examined, and pooled estimates were recalculated as needed to assess their impact. Together, these procedures ensured that the findings were robust, not driven by individual studies, and provided a reliable basis for interpreting prevalence estimates in a public health context.

## 3. Results

### 3.1 Study Selection

Figure 1 illustrates the systematic selection and screening process of studies included in the review.

**Fig. 1.**
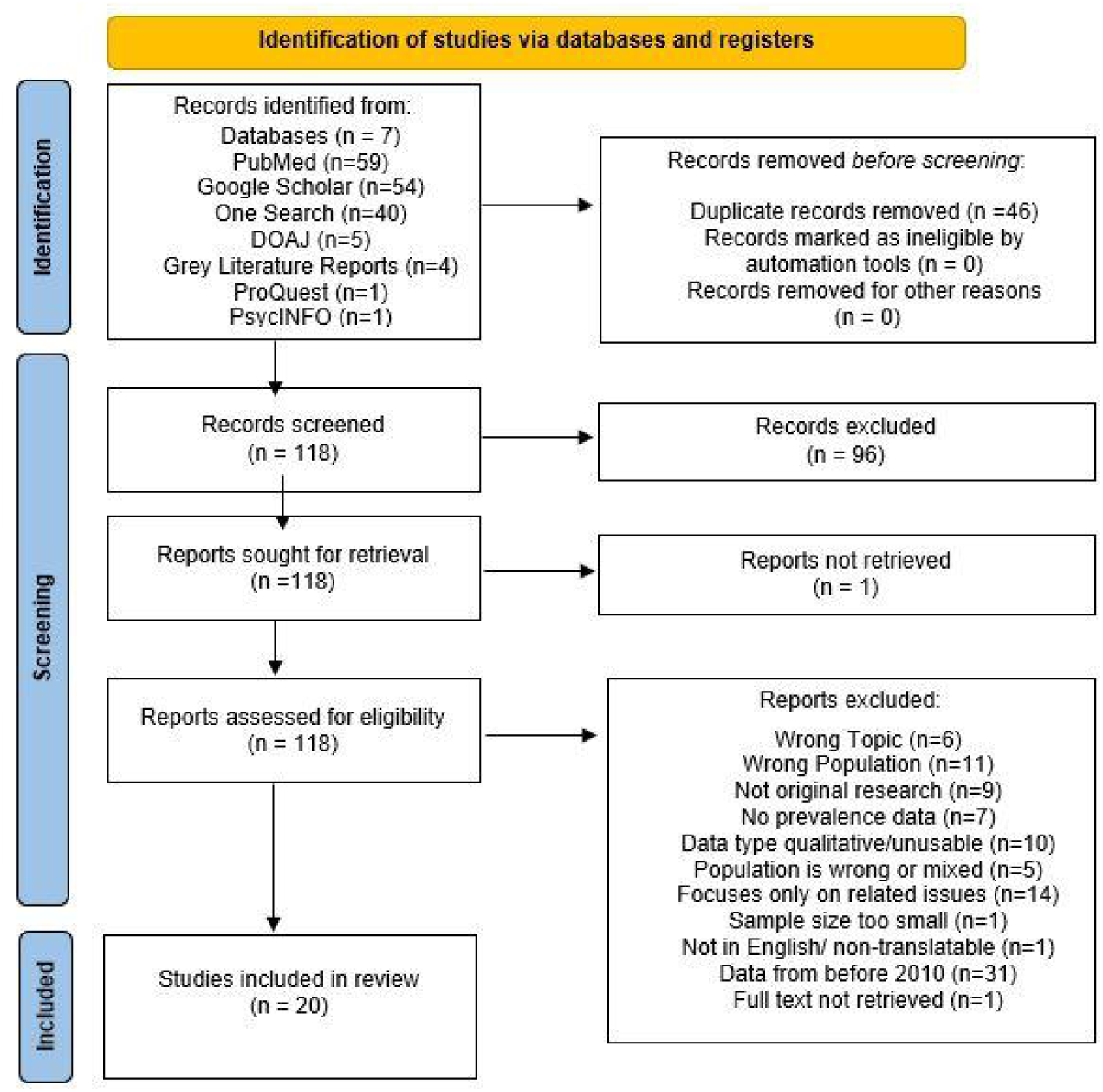
Prisma flow chart. Created according to PRISMA 2020 guidelines, using the SHINY flow diagram creation tool. The identification process included database searches and the elimination of duplicate titles by research assistants. A total of eleven exclusion criteria were applied to the remaining 118 studies during the screening process.

### 3.2 Study Characteristics

The overall prevalence analysis included 20 studies with 4,871 participants, while the 12-month analysis included 14 studies with 4,693 participants. All studies were published after 2010 and were primarily observational, using cross-sectional survey designs. For the overall prevalence analysis, removing any one of the 20 studies resulted in pooled estimates that remained stable within a narrow range of 0.28 to 0.37. The high heterogeneity also remained consistent at approximately 96–97%, confirming that no individual study was responsible for the findings. The 12-month prevalence analysis also showed that the pooled estimate remained stable during its leave-one-out analysis.

The characteristics of the study populations varied considerably. Individual sample sizes ranged from 34 to 1,115 participants. While many studies surveyed a general population of homeless women, several focused on specific high-risk subgroups, including women with disabilities, mental health conditions, LGBTQ+ women, and HIV-positive women. The systematic assessment concluded that the findings are robust, as they are not significantly skewed by publication bias or the undue influence of any single study.

A key source of variation across studies was the operationalization of sexual assault. Definitions ranged from narrow measures, such as completed rape, to broader concepts, including survival sex, sexual coercion, or any unwanted sexual contact. Furthermore, recall periods differed, with some studies assessing lifetime exposure while others focused on events within the last 12 months, 6 months, or 30 days, necessitating separate syntheses.

### 3.3 Risk of Bias in Studies

Another critical aspect of the discussion is the potential for methodological bias among the included studies. We used Egger’s test to assess the presence of small-study effects, which can signal publication bias or inflated effect sizes in smaller studies. For the overall prevalence analysis (k = 20), Egger’s test showed no significant small-study effect (t = 0.48, df = 20, p = 0.639). For the 12-month prevalence analysis (k = 14), Egger’s test similarly showed no evidence of small-study effects (p = 0.7308).

However, it is essential to note that Egger’s test has limited statistical power when the number of included studies is small (in this case, k = 20), so the absence of significant findings should be interpreted with caution. Based on this stability, the findings are considered moderately reliable, as indicated by the ICC (Intraclass Correlation Coefficient), suggesting they exhibit consistent patterns but require further investigation in studies aiming for reproducibility.

A leave-one-out procedure revealed that removing any single study didn’t significantly alter the overall results; the pooled estimates remained within a narrow range (0.28–0.37), and I² remained stable at approximately 96–97%. Influence appeared to be distributed across the dataset, which is consistent with the overall variability in effect sizes.

### 3.4 Results of Individual Studies

The characteristics of the 20 studies included in the overall prevalence meta-analysis are detailed in Table 1. The studies varied considerably in terms of the definition of sexual assault, population characteristics, sample size, and pooled prevalence. As shown, individual prevalence rates reported in the studies ranged widely, from as low as 2.6% in a study of HIV-positive women to as high as 92.3% in studies focusing on women with disabilities, highlighting the significant between-study heterogeneity that was formally assessed in the synthesis.

### 3.5 Results of Syntheses

Across 20 studies, the overall prevalence of sexual assault among homeless women was calculated to be 39%. Using a random-effects model with logit transformation, the confidence interval for this estimate was 25% to 56% (see Table 1).

For comparison, we also calculated a common-effect estimate of 32.7% (95% CI 0.314– 0.341) to contrast with the random-effects model, which accounts for variation across studies. These values come directly from the R output.

The pooled estimated 12-month prevalence was 22%. In practical terms, this indicates that more than one in five homeless women experienced sexual assault within a single year. This result highlights the ongoing and serious risk of violence faced by this population.

Among the groups and subgroups analyzed, women with disabilities had the highest reported prevalence of sexual assault, with one study showing over 92% of women affected. In contrast, the lowest prevalence was found among HIV-positive women, where only 2.6% reported sexual assault, though this finding was based on minimal data. Elevated rates were also seen among LGBTQ+ women (33%) and women with mental health conditions (33.6%).

The interpretation of these subgroup findings is constrained by small sample sizes, which limit reliability and weaken the conclusions that can be drawn. This means the study results vary significantly. This could be due to studies that examined different subgroups and were conducted in various geographic regions. When heterogeneity was low, a fixed-effects model was used; when it was moderate to high, a random-effects model was employed.

To identify sources of heterogeneity and disentangle subgroup effects, subgroup analyses and meta-regression analyses were conducted. Publication bias was analyzed with funnel plots and Egger’s test. This heterogeneity likely reflects differences in study populations across different subgroups and geographic areas.

Between-study heterogeneity was very high (Q = 732.24, df = 19, p < 0.0001; I² = 97.2%, 95% CI 96.4–97.7; τ² = 1.95). This pattern suggests that the observed variation is unlikely to be due solely to chance. Instead, it likely shows real, statistically significant differences in risk. These differences appeared across various factors, including population traits like disability status, HIV status, and LGBTQ+ identity. Variation was observed across different geographic locations and in how studies defined and measured sexual assault.

**Fig. 2.**
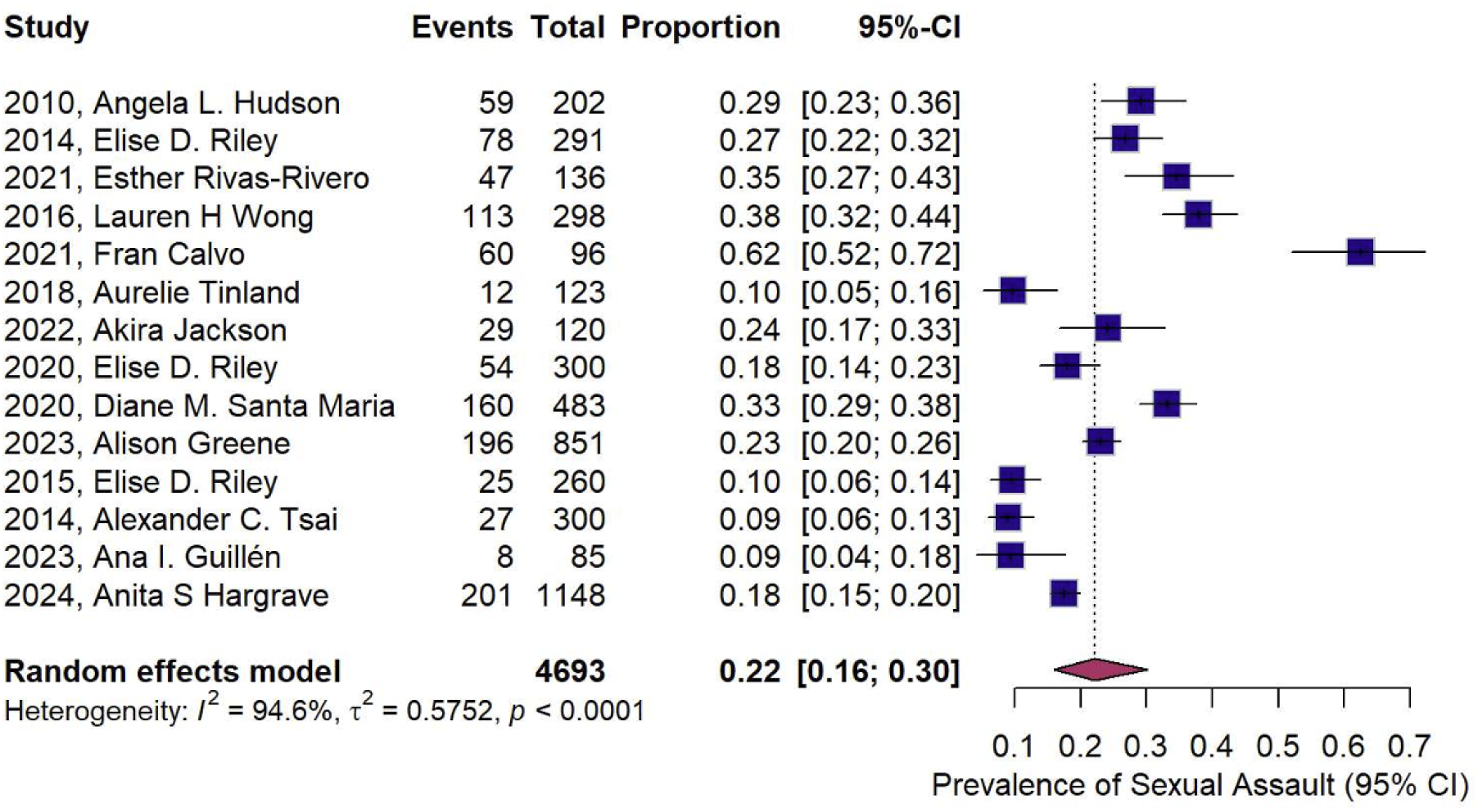
Forest Plot of 12-Month Prevalence of Sexual Assault. This figure shows the prevalence of each of the 14 studies included in the 12-month analysis, with a pooled prevalence of 22% among 4,693 total subjects. Sample sizes ranged from 85 to 1148, with case counts ranging from 8 to 201.

**Fig. 3.**
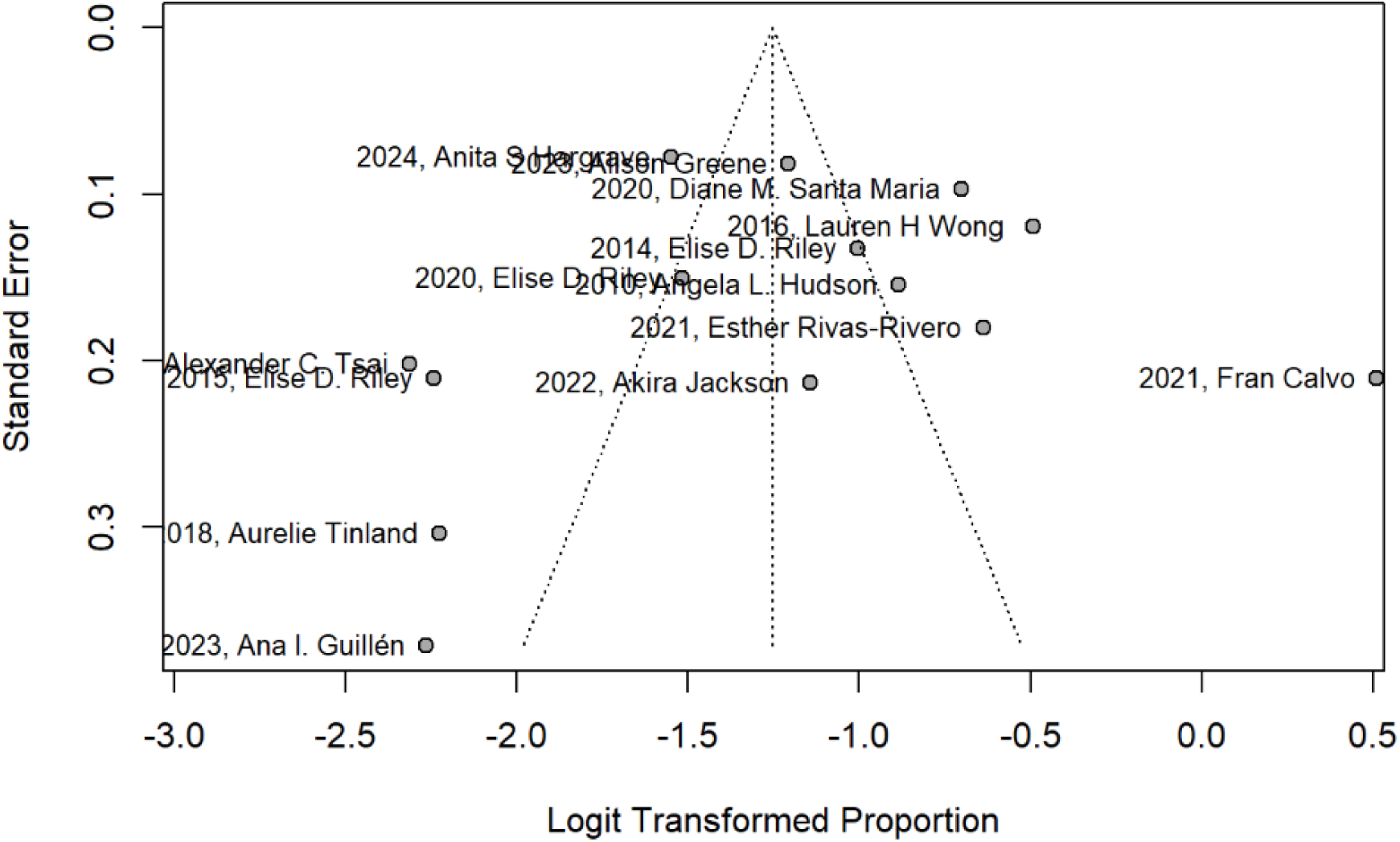
Funnel Plot for 12-Month Prevalence Synthesis. Egger’s regression tests, graphed as a funnel chart. This chart assesses overall effect sizes for the 14 studies included in the 12-month prevalence synthesis. Egger’s test similarly found no statistically significant evidence of funnel plot asymmetry (p = 0.7308).

Subgroup analyses demonstrated significant differences. For instance, the chi-squared test discovered significant variation between the subgroups, highlighting that being a member of a subgroup as a homeless woman could confer a greater risk of sexual assault, and that the various subgroups had differing risk levels and factors. (χ² = 152.3, df = 6, p < 0.0001; Figure 4).

**Fig. 4.**
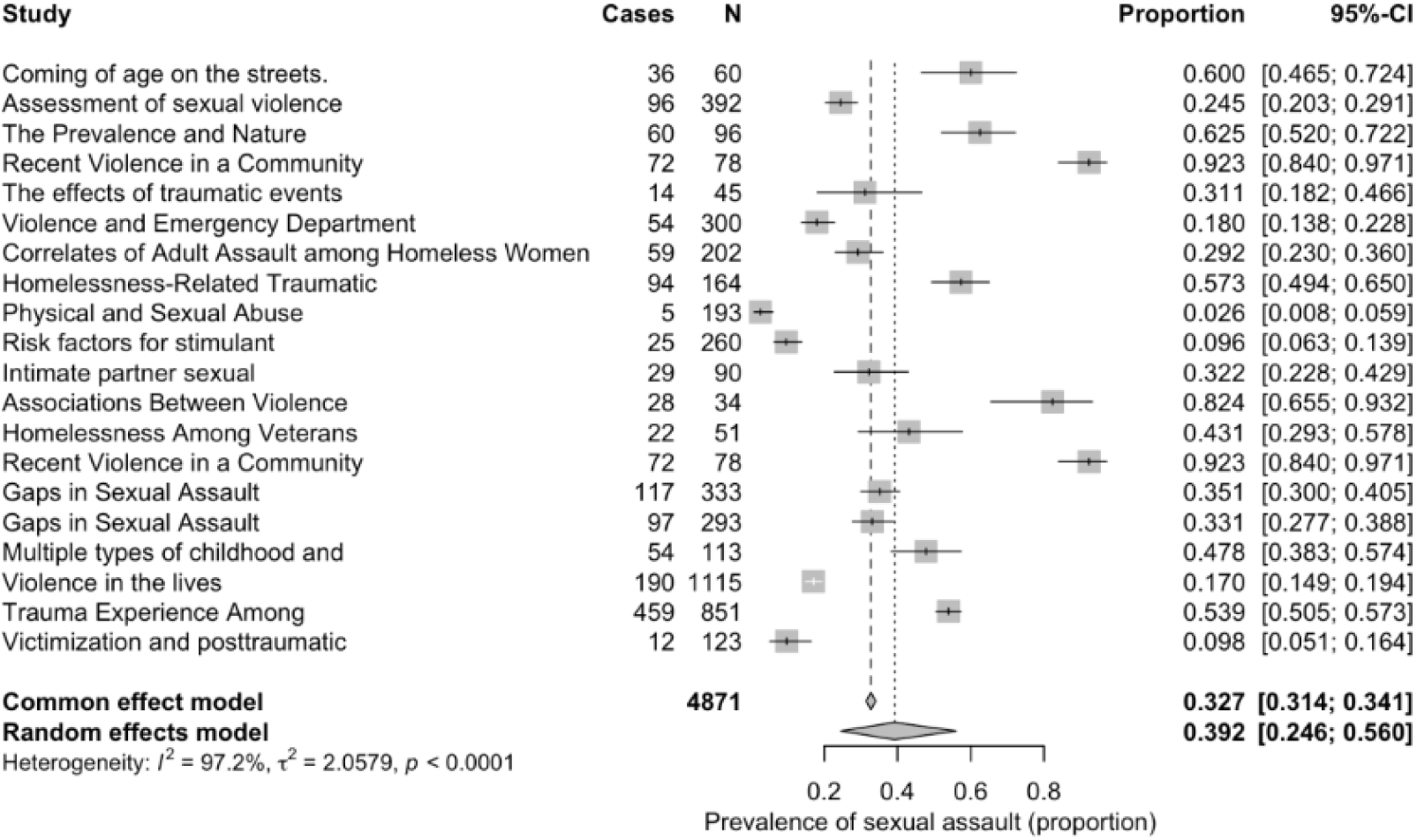
Overall forest plot for lifetime analysis. Each of the 20 studies used both the standard and random effects models, with a heterogeneity of 97.2% observed. The contribution of each study to the overall effects was estimated, with 95% confidence intervals (CIs). The average contribution to the values was 32.7% within the common-effects model and 39.2% within the random-effects model. The sample size from the studies had an extensive range, from (n =34 −1115), and a case range of 5- 459.

To further explore potential sources of heterogeneity, we conducted a meta-regression using study quality score and overall risk of bias as moderators. Both variables were statistically significant predictors of variability in effect size. Higher study quality was associated with smaller effect sizes, whereas a higher risk of bias was associated with larger effect sizes. Together, these moderators explained a meaningful proposal.

### 3.6 Reporting Biases

An assessment for reporting bias was conducted to evaluate the potential for small-study effects across both the lifetime and 12-month prevalence syntheses. For the overall prevalence analysis (k=20), Egger’s regression test found no significant evidence of funnel plot asymmetry (t = 0.48, df = 20, p = 0.639), suggesting an absence of small-study effects. Visual inspection of the funnel plot confirmed broad symmetry, supporting this statistical finding. To further test the robustness of this conclusion, a Duval and Tweedie trim-and-fill analysis was performed, which imputed two hypothetical studies at the low-prevalence end of the plot. This adjustment yielded a slightly lower pooled estimate of 0.33 (95% CI: 0.20–0.50), while heterogeneity remained essentially unchanged (I² = 97.3%).

For the 12-month prevalence analysis (k = 14), Egger’s test similarly found no statistically significant evidence of funnel plot asymmetry (p = 0.7308). It is important to interpret these results with caution for both analyses. Egger’s test has limited statistical power when the number of included studies is small and heterogeneity is high, as was the case in both syntheses. Nonetheless, the combined findings suggest that potential publication bias does not alter the substantive conclusion that there is an elevated prevalence of sexual assault among homeless women, reflecting the overall robustness of the results.

Egger’s regression test, which assesses publication bias due to small-study effects, indicated little evidence of small-study effects (p = 0.639). Visual inspection of the funnel plot (Figure 5).

**Fig. 5.**
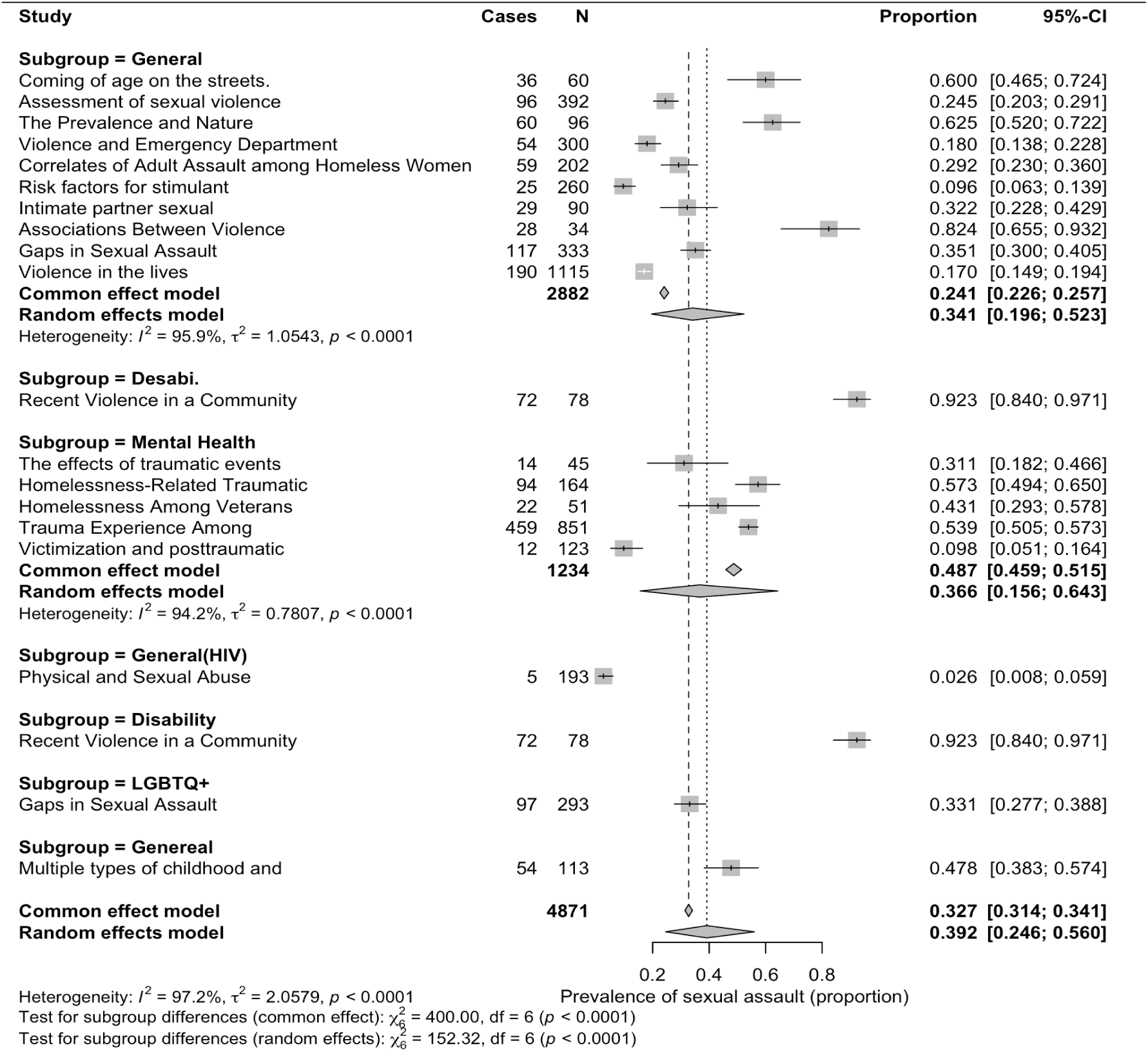
Forest plot for lifetime analysis. Each of the 20 titles was sorted by subgroup (LGBTQ+, mentally ill, disabled, HIV positive), with an analysis of the proportion of prevalence of sexual assault by group. Data analysis was conducted in R, and heterogeneity, common effects, and random effects were used to test for subgroup differences. The between-study heterogeneity partially accounted for the extremely high I² value (∼97%).

**Fig. 6.**
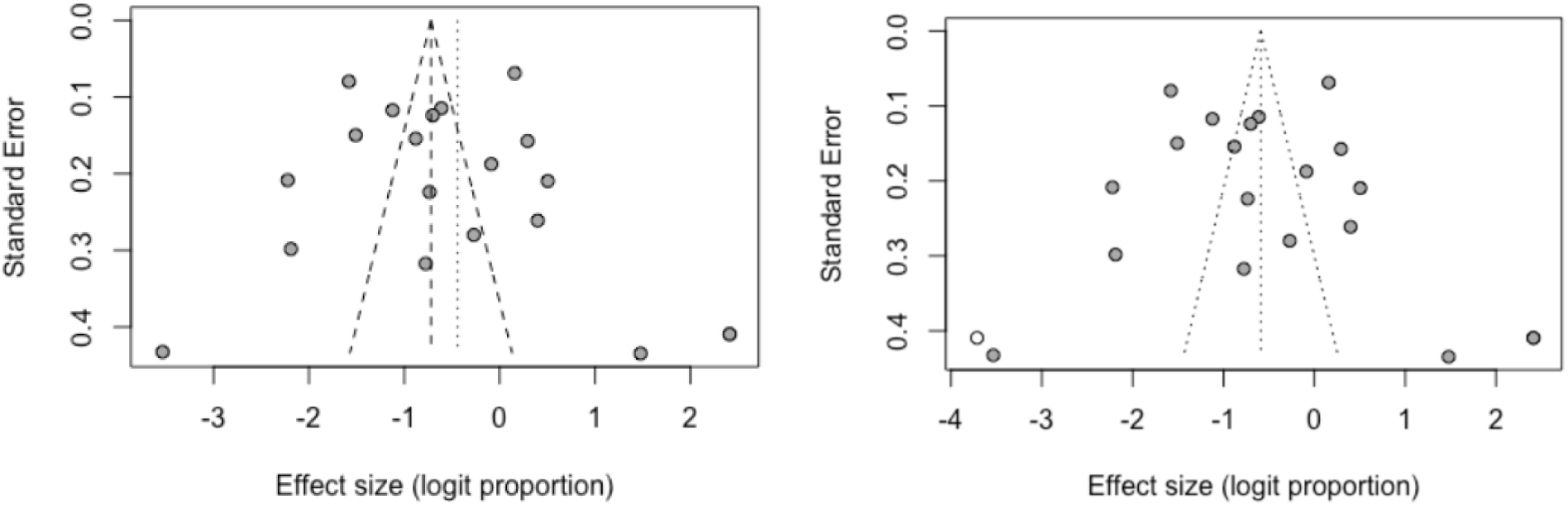
Egger’s regression tests, graphed as a funnel chart. The right chart assesses overall effect sizes, and the left analyses small-study effects, performed using R software, confirming broad symmetry and suggesting that the results are representative of the overall state of research in this area. Trim-and-fill analysis imputed two studies at the low-prevalence end, yielding an adjusted pooled estimate of 0.33 with a 95% CI of 20-50%. Heterogeneity remained largely unchanged (I² = 97.2%; Q = 851.52, df = 23). These findings suggest that potential publication bias likely does not alter the substantive conclusion of elevated prevalence, reflecting the overall robustness of the results.

The systematic assessment revealed robust findings, with no evidence of significant publication bias or undue influence from individual studies. For the overall prevalence analysis, removing any one of the 20 studies resulted in pooled estimates that remained stable within a narrow range of 0.25 to 0.39. In the lifetime analysis, high heterogeneity remained consistent at approximately 96–97% (I² = 97.2%), confirming that no individual study was responsible for the findings.

The results display moderate reliability, as indicated by the IIC (Intraclass Correlation Coefficient). The moderate reliability score suggests that the findings remain stable over time, exhibiting consistent patterns across prevalence estimates, but suggesting moderate to low replicability across studies. In the lifetime analysis and subgroup syntheses, Baujat plots indicate that heterogeneity was distributed across several studies, with only slight skewing toward studies reporting extreme prevalence values and toward those with larger sample sizes. (e.g., ∼92% in the disability-focused sample and ∼2.6% in HIV-positive samples).

Additional influence tests (studentized residuals, DFFITS, Cook’s distance, leverage, and τ² deletion) confirmed that no single study exerted a disproportionate effect on the results. Together, these sensitivity Analyses suggest that the observed heterogeneity is not due to outliers but rather to genuine variation across study populations and methodological contexts. The Baujat plot (Figure 7) shows that heterogeneity was distributed across several studies, with trauma experience and violence in the lives affected the most. The leave-one-out sensitivity analysis (Figure 8) confirmed that removing any single study did not substantially alter the pooled prevalence, which remained between 0.28 and 0.37.

**Fig. 7.**
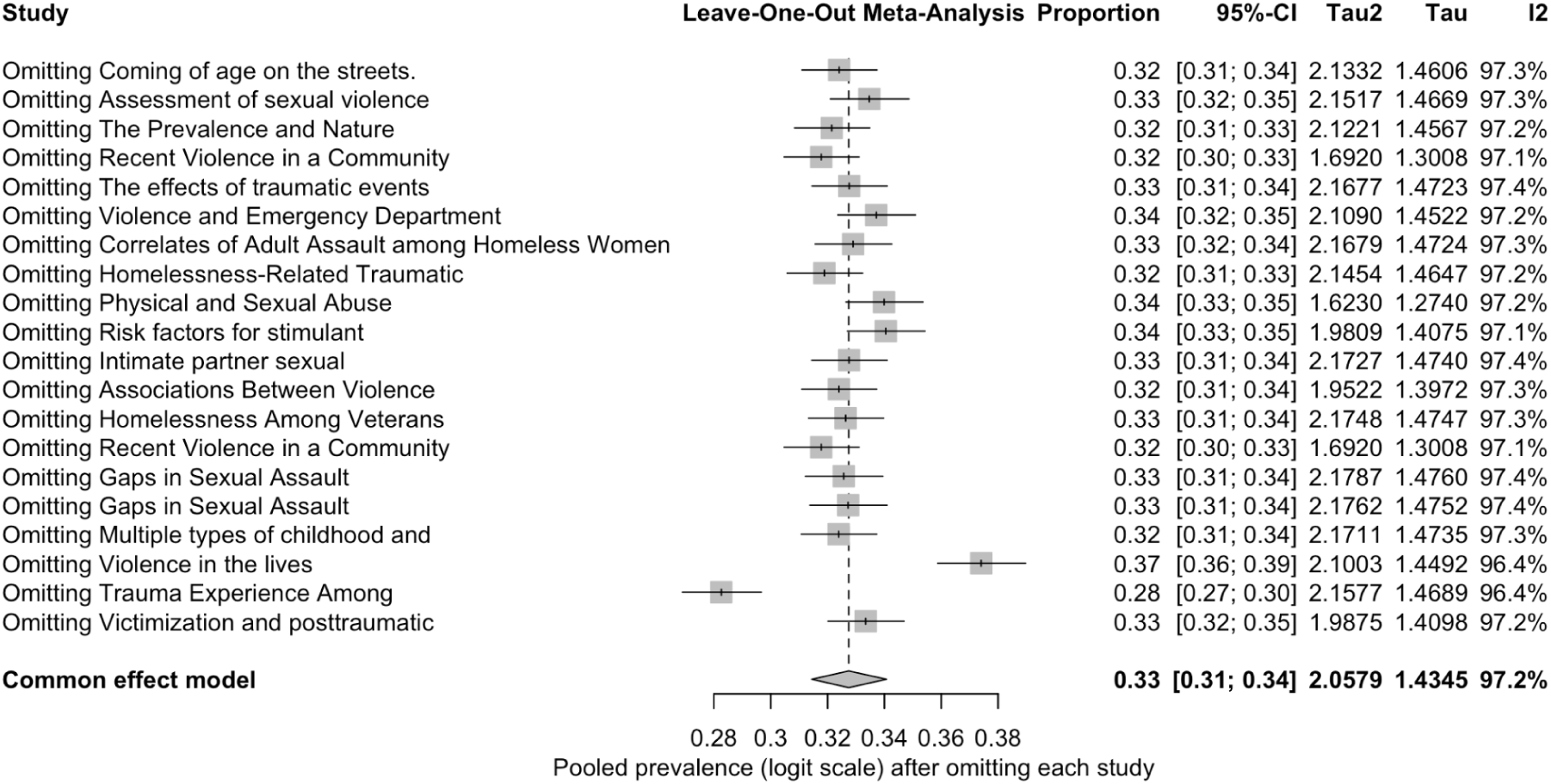
Leave-one-out Meta-Analysis Logit Proportion, with every study omitted once.Figure 7 presents the results of the leave-one-out meta-analysis, illustrating the influence of each individual study on the overall pooled logit proportion. This analysis assesses the robustness and stability of the meta-analytic findings by sequentially omitting each study in turn and recalculating the combined effect size.

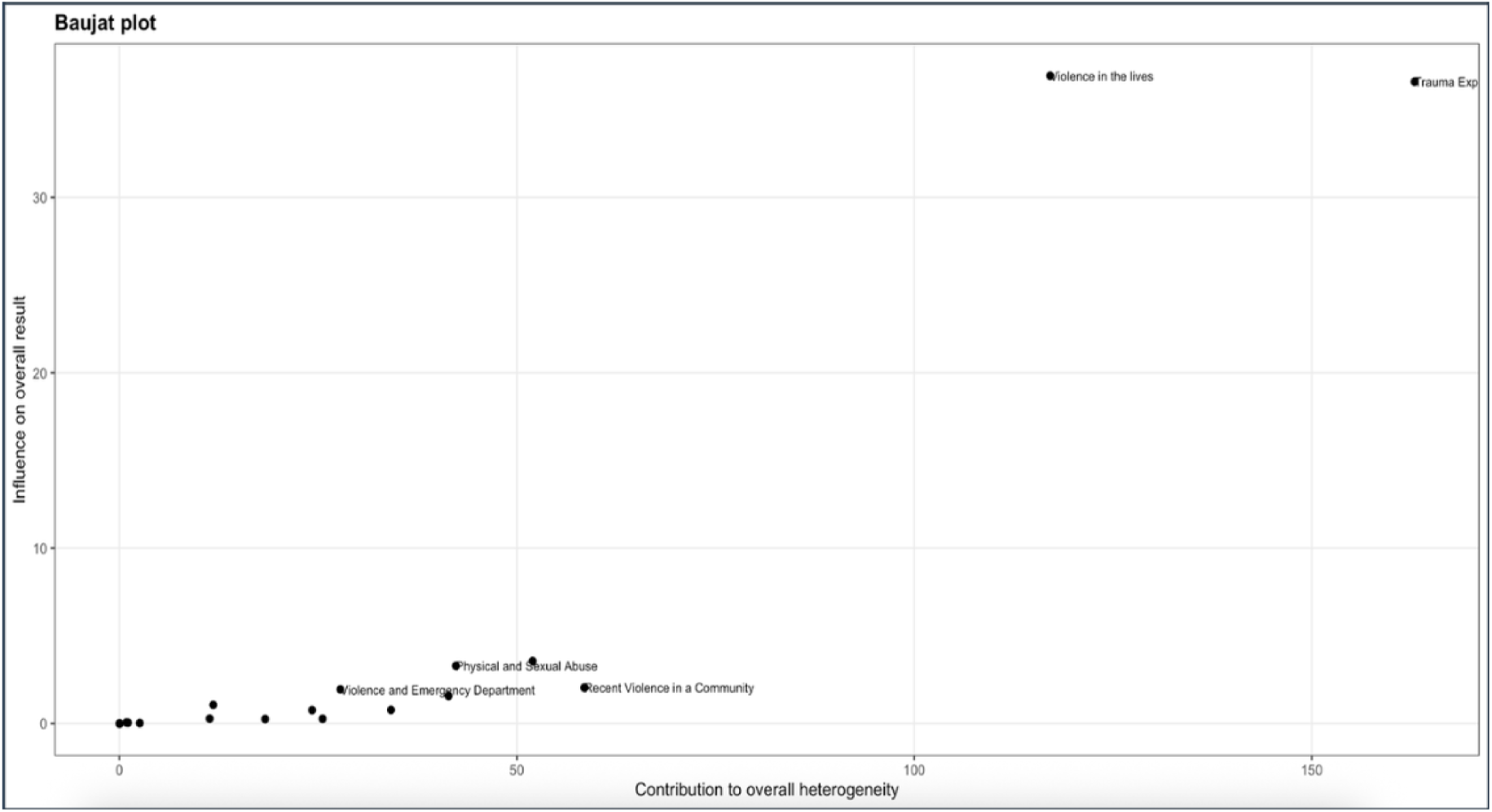
Baujat Plot (Fig. 8.Lifetime Analysis). The Baujat plot identifies two studies; *Trauma Exposure* and *Violence in the Lives*, as clear outliers positioned in the upper-right quadrant (Quadrant I). These studies exhibit a disproportionately high contribution to the overall heterogeneity (x-axis) and exert the greatest influence on the pooled effect size (y-axis). In contrast, the remaining studies, including *Physical and Sexual Abuse*, *Violence and Emergency Department*, and *Recent Violence in a Community*, are clustered near the origin (0, 0), indicating minimal contribution to both heterogeneity and the overall summary estimate. Their proximity to the origin suggests consistency with the pooled effect, implying negligible influence on the observed between-study variability. Overall, this pattern indicates that the pooled estimate is robust, with heterogeneity largely driven by a small subset of influential studies.

**Fig. 9.**
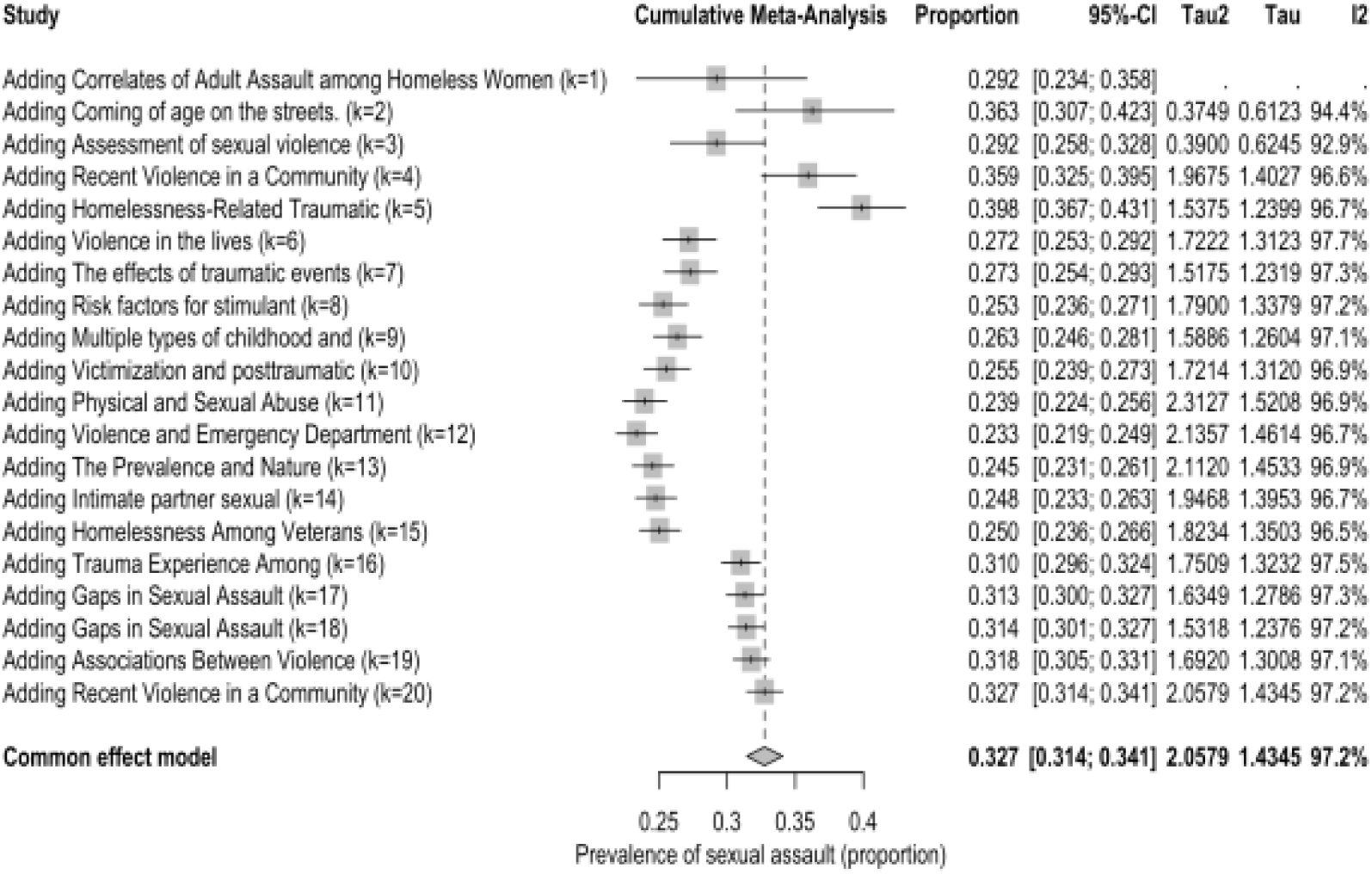
Cumulative Meta-Analysis by Year (Stability over time). Fig. 9 represents a cumulative meta-analysis by year of prevalence of sexual assault across the 20 studies included. The figure indicates a pooled prevalence of 0.327 with a 95% CI ranging from 0.314-0.341.

**Fig. 10.**
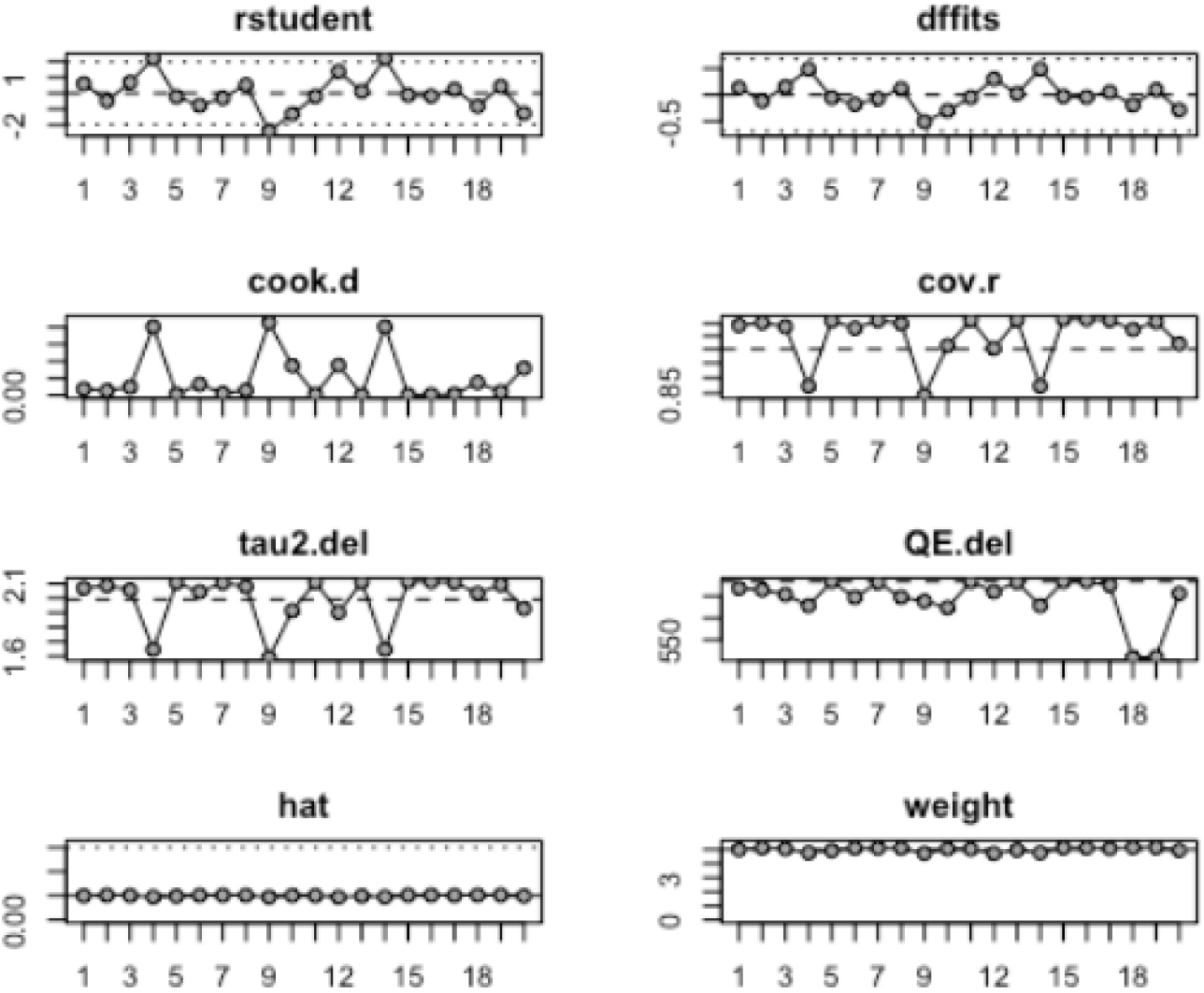
Other influence diagnostics. Performed via ‘metafor’ for regression-based meta. Studentized Residuals (rstudent)for each observation. A significant absolute value greater than 2 or 3, suggests a large residual that is not due to leverage alone and may indicate an outlier study. The DFFITS plot displays the value representing the change in the predicted value when a point is deleted. Points with large DFFITS values (positive or negative) indicate a strong influence on the model’s fit. Cook’s Differentials (cook.d) were computed to measure the influence of each data point on the entire model. Points exceeding 0.5 are potential influential points worth further examination. Covariance ratio (cov.r) measures the covariance matrix of coefficients if a point is removed. A value deviating from 1 shows that a study has a greater influence on the overall result. Three studies in the Covariance ratio (cov.r) graph indicate greater effects. τ²_deleted (tau2.del) quantifies the influence of an observation on the model’s variance components. A high value of τ²_deleted suggests that the observation has a considerable impact on the variance estimates. Quasi-residual (QE.del) for each observation is used to identify outliers. A considerable absolute value of the quasi-residual indicates that the observation is an outlier, even after accounting for its leverage. The plot of hat values (hat) identifies high-leverage points. Studies with large hat values have unusual predictor variable values, making them influential on the regression line regardless of their residual. The (weight) graph represents the standardized weights for each observation.

The 12-month prevalence analysis also indicated moderate reliability. Egger’s test (p = 0.7308) showed no evidence of significant publication bias, and a leave-one-out procedure demonstrated that removing any single study did not substantially change the pooled prevalence (which remained around 22%).

Although heterogeneity was high (I² ≈ 94.6%), influence diagnostics (studentized residuals, DFFITS, Cook’s distance, leverage, and τ² deletion) confirmed that any one study did not drive the findings. Unlike the lifetime analysis, fewer subgroup data were available for the 12-month recall period, limiting diagnostic detail. Nonetheless, the stability of the pooled estimates across multiple checks supports the conclusion that results reflect consistent patterns across studies.

## 4. Discussion

### 4.1 Summary of Evidence

The results of this study confirm that homeless women face alarmingly high rates of sexual assault. Homeless women living in the United States are found to have an overall prevalence of sexual assault of 39%, compared to the 19.3% prevalence among all women in the United States [2]. However, the risk of sexual assault does not appear to be evenly distributed across all groups of women experiencing homelessness [3]. These findings are based on a total pooled sample of over 4,871 women experiencing homelessness across the included studies. To evaluate the overall prevalence of sexual assault among homeless women, both common-effect and random-effects models are utilized. The common-effect model assumes that all studies estimate a single, shared risk of sexual assault across all subgroups, thereby offering a broad overview for comparison with the random-effects model.

The random-effects model, in contrast, highlights the prevalence of sexual assault across the subgroups. Therefore, the random-effects model was employed more extensively in the data analysis, given the diversity of risk factors and experiences among women experiencing homelessness. The heterogeneity of subgroup sexual assault prevalence strongly supports the use of the random-effects model as the more appropriate framework for interpreting the overall findings. The random-effects model also accounts for variation between studies, including location, sample size, and subgroups surveyed.

Our subgroup analysis revealed significant differences between populations (χ² = 152.32, df = 6, p < 0.0001), with prevalence rates ranging from approximately 2.6% among HIV-positive women (n = 78) to 92.3% among women with disabilities. In comparison, women with mental health conditions showed rates between 31.1% to 57.3% across multiple studies (combined n = 1,234)^(1)^. LGBTQ+ individuals also experienced elevated rates, with a prevalence of 33.1% (n = 293) [14].

The subgroup analysis revealed significant differences, indicating that the varying rates across subgroups are unlikely due to chance. The most striking result came from one study focused on women with disabilities, where the prevalence of sexual assault was very high, limiting the replicability and statistical significance for the disabled subgroup. Other groups, including LGBTQ+ individuals and those with mental health challenges, had moderate prevalence of assault, but there was significant variation within those subgroups^(4)^.

Women with disabilities face the highest documented risk of sexual assault, according to the literature review. Our findings align with previous research on the general, housed population. People with disabilities who are stably housed face far greater rates of sexual assault and domestic abuse than the general population. 83% of intellectually disabled women will be assaulted in their lifetime, compared to 26.8% of non-disabled women, and 2 out of 5 female rape victims are disabled women. Over half of the disabled women surveyed from 2011-2017 by the National Survey of Family Growth experienced sexual assault [15]. All people with disabilities reported sexual assault rates of 60% over their lifetime, meaning the risk of sexual assault is 3 times higher than in the general population [2]. One factor that immediately puts disabled people at risk is their physical or mental disability itself, which leads to perpetrators assuming they will not fight back or tell anyone.

Another study found that among women aged 18–44 years with sensory, physical, or cognitive disabilities, close to 30% reported experiencing forced VI at least once in their lives, significantly more than the prevalence among nondisabled women, consistent with previous studies.5,13,14,32,33 For those with multiple disabilities, the prevalence was >40%, notably on the upper range of previous estimates.13,14 In addition, 34.2% of women with multiple disabilities experienced either physical/nonphysical force during their first VI, twice the prevalence among nondisabled women (17.7%). After controlling for confounders, women with any disability type included in this study are significantly more likely to have experienced forced sex during their lifetime than nondisabled women, with the most significant risk among women with ≥2 disabilities [15].

Mentally disabled or developmentally different adults may not be given proper sex education or may not be educated on consent, because it is assumed they will never use the information. Another factor to consider is abuse by caregivers, relatives, or peers before becoming homeless. Caregivers or those who have immediate access to the disabled person may threaten or coerce them through threatening their well-being or using their disability as a weapon, which is particularly common when a disabled person has a sole caregiver or is placed in a group living home. People with disabilities may also face ableism. They may therefore be less likely to be believed if they attempt to report an assault, adding another layer of difficulty in receiving help as a victim and accessing accurate rates of assault for disabled homeless women [16].

Similarly to disabled women, mentally ill women are at a higher risk of sexual assault than other subgroups. 31.1-57.3% of mentally ill homeless women faced sexual assault over their lifetime, which is also significantly higher than the prevalence rates among the general population. Stably housed women with mental illness also display a much greater risk of victimization and sexual assault. When studying forensic reports of 7,455 sexual assaults, it was found that the rate of sexual assault of mentally ill individuals was higher, and that perpetrators are less likely to be romantic partners, and are more likely to be teachers, colleagues, peers, or family members. This suggests that, similar to the disabled subgroup, mentally ill victims may be more vulnerable due to caretakers or peers believing that they will not be considered or will be incapable of defending themselves. Mentally ill victims also had a 60% rate of prior victimization compared to 40% of the general population, with more prior sexual violence occurring under the age of 14 in the mentally ill group. There was also significant overlap of mental illness being reported and other factors, such as blindness, severe psychosis, drug usage, and other physical and cognitive disabilities. This study also specified that mentally ill victims are less likely to have a permanent address (25.6% vs 19.2%; p = 0.031) [17].

Another group of women who may face structural and social barriers to preventing assault are LGBTQ+ women. LGBTQ+ women experiencing homelessness often face multiple layers of discrimination. They might be denied access to safe, gender-affirming shelters or find themselves in openly hostile environments. This forces many women into unsafe situations, such as sleeping outdoors or relying on unsafe housing arrangements, where the risk of sexual violence rises.

In contrast, HIV-positive women demonstrated a notably low prevalence estimate (2.6%). This result should be interpreted cautiously, given the small sample size and the potential for under-reporting or sampling bias.

The pooled prevalence among homeless women (39%) was nearly double the national estimate for all U.S. women (20%, NISVS). Subgroup findings align with prior research: women with mental health conditions (36.6%) and LGBTQ+ women (33.1%) showed elevated prevalence, women with disabilities faced extreme vulnerability (>90%, though based on a single study), and HIV-positive women reported the lowest prevalence (2.6%). Overall, these results emphasize both the significant burden of sexual assault among homeless women and the variability across subgroups [18].

Overall, these patterns show how multiple forms of marginalization—whether based on ability, mental illness, gender/sexual identity, or health status—can overlap and increase a person’s risk of experiencing violence. To reduce sexual assault among women experiencing homelessness, these differences must be understood and addressed, rather than treating the population as if they are a uniform group.

Based on the study conducted, the prevalence of sexual assault among homeless women observed in this study is substantially higher than in the general population. National data from the National Intimate Partner and Sexual Violence Survey (NISVS) estimates that approximately 19.3% of U.S. women report lifetime sexual assault or attempted rape. In contrast, our pooled prevalence among homeless women was 39%. [18]. Subgroup-specific findings also align with and expand upon prior research, with women with mental health conditions (36.6%) and LGBTQ+ women (33.1%) showing consistently elevated prevalence, while women with disabilities demonstrated extreme vulnerability with rates exceeding 90%, and HIV-positive women reported the lowest prevalence at 2.6%. The rates in our study are consistently higher across most subgroups. The pooled prevalence across studies was nearly double the national baseline. Collectively, these results emphasize the significant burden of sexual assault among homeless women, as well as the variability in risk across different subgroups.

A key methodological finding of this meta-analysis was the exceptionally high degree of heterogeneity among the included studies (I² ≈ 97.3% (95% CI 96.6–97.8) and Q = 739.93 (df = 20, p < 0.0001). This variability likely reflects fundamental differences in populations, settings, and study designs, including whether participants had disabilities, whether samples were clinical or community-based, and how sexual assault was defined. Such heterogeneity is expected in diverse populations and supports the choice of a random-effects model.

Sensitivity and influence analyses indicated stable results. Leave-one-out procedures showed that removing any single study did not materially alter the pooled prevalence (range, 0.32–0.37), and I² consistently remained high (I² = 97%, Q = 851.52, df = 23). A Baujat plot identified studies with extreme prevalence estimates (e.g., disability-focused and HIV-positive samples) as contributing most to heterogeneity, along with larger studies that naturally carried more weight. However, diagnostic testing using Cook’s distance, DFFITS, and studentized residuals did not flag any study as disproportionately influential, suggesting that variability was broadly distributed.

Publication bias was assessed using Egger’s test, which found no evidence of small-study effects (t = 0.44, df = 18, p = 0.6670). This test has limited statistical power due to the modest number of studies (k = 20).

Together, these methodological tests indicate high heterogeneity between studies, without overall assault rates for homeless women being driven by small-study effects, single studies, or extreme values. The subgroup findings were limited by the small portion of studies, notably for the disabled, pregnant, and LGBTQ subgroups. The findings should therefore be interpreted as preliminary, and appropriate caution should be taken; these results are not generalizable for all subgroups. This meta-analysis is invaluable for establishing a framework for variability across study populations and for highlighting which risk factors are understudied. The prevalence number across all studies does highlight a key finding that describes homeless women’s risk of sexual assault.

This research presents a preliminary hypothesis that certain homeless women are at greater risk of assault and may be less likely to be studied. The results point to sexual assault risk not being evenly distributed among homeless women. Further literature review has suggested that there may be less reporting and research of assault directed at disabled women. Pregnant women in homeless settings were hypothesized to be at greater risk of assault due to vulnerability. Still, the data analysis did not provide significant findings on the prevalence of sexual assault among pregnant women, due to a lack of distinction between pregnant and non-pregnant women in most studies.

### 4.2 Limitations

This study has several significant limitations. First, heterogeneity was notably high (I² ≈ 97%), reflecting differences in study subgroups, geographic locations, designs, and time periods. While random-effects models account for such variation, the pooled prevalence should be interpreted as an approximate range rather than an exact value.

Furthermore, subgroup analyses are limited by data availability. Disability estimates, though strikingly high, are based on only two studies. The HIV and LGBTQ+ subgroups also came from a single study. Pregnant women, transgender women, and sex workers are populations hypothesized to be highly vulnerable, and they are not represented. This underrepresentation limits precision and the ability to generalize about subgroup findings.

It is crucial to note that geographic diversity was heavily skewed. Nearly all studies were conducted in the United States, with only two from Europe (France and Spain). Findings may therefore not reflect prevalence patterns in regions with different cultural, social, or policy contexts.

It should also be noted that study designs are overwhelmingly cross-sectional, which limits the ability to establish causal pathways or changes over time. Additionally, definitions of sexual assault are inconsistent, with some studies restricting their definition to completed rape while others include broader forms of sexual violence. The definition used by this meta-analysis was “forced penetration (rape), unwanted sexual contact, coercion through threats, intimidation, or manipulation,” and all included studies had a definition that was similar to the one used by this study.

Finally, underreporting is hypothesized to be highly likely. Sexual assault is thought to be widely underreported due to stigma, fear of retaliation, or mistrust of institutions. Among homeless women, these barriers may be even more substantial due to financial vulnerability, meaning actual prevalence is probably higher than observed. Moreover, several studies recruited participants through shelters or clinics, leaving unsheltered women, who may face greater risks, underrepresented.

### 4.3 Implications for Practice

Policymakers should take note of the heightened vulnerability of homeless women to sexual assault and consider legislative interventions and practical solutions. Sexual assault is a complex phenomenon with no one-size-fits-all solution. However, there are practical steps that policymakers can take to address the alarming rates of violence and coercion captured in the surveyed studies.

It is well known that rape is an underreported crime, with some research suggesting that at least one million rapes are left out of crime statistics annually [19]. One of the reasons for this phenomenon was also a significant challenge and limitation of this study: the necessity of navigating substantially different definitions of “rape” and “sexual assault” in the academic literature on this topic.

While attitudes towards sexual assault have evolved, legal definitions of “rape” broadly continue to exclude economic coercion or exploitation. In addition, criminalization of prostitution risks compounding the injury to women who suffer such exploitation. Legislators should consider decriminalization of sex work or, at a minimum, mandating the consideration of economic exploitation as a mitigating factor in sentencing decisions.

Future legislative and policy interventions should recognize that poverty, economic desperation, and sexual assault are intertwined in ways that have not previously motivated efforts to ameliorate the problem [20]. The results of this study suggest that policy interventions aimed at improving the socio-economic status of women at risk for sexual assault, particularly those aimed at providing stable housing, are likely to have a significant impact. While the design of this study does not allow for definitive conclusions on the causative relationship between sexual assault and homelessness, it does provide some evidence that such a relationship exists.

Some research has found that while temporary protective orders leave women at greater risk for gender-based violence, permanent civil protective orders measurably decrease the incidence of such violence [9]. This suggests that judges’ attitudes and sensitivities towards the issue are an essential factor in the ultimate efficacy of legal interventions. Policymakers should consider making education on these issues mandatory for judges and magistrates who issue civil protective orders.

In addition, more consistent enforcement of civil protective orders provides a heightened level of protection for women at risk of sexual assault. Police departments should be encouraged to robustly enforce civil protective orders and encourage victims to apply for them. This solution has the advantage of avoiding reliance on the legislative process, which can often be slow and inefficient.

Finally, the subgroup analyses discussed above suggest that any of these interventions are most urgently needed for LGBTQ+ women and women with disabilities.

### 4.4 Implications for Research

Although this meta-analysis supports that sexual assault is highly prevalent among homeless women, significant gaps remain. Sex workers, groups consistently identified as high-risk in non-homeless literature, are almost absent from included studies. The lack of findings focused on sex workers underscores the need for deliberate recruitment of these populations in future research.

The disability subgroup, which had the highest prevalence (over 90%), was based on a single study. Similarly, the HIV positive subgroup demonstrated markedly low, but only drew upon one study for its prevalence data. Additional research is needed to verify these extreme estimates, identify the mechanisms underlying differences in risk, and determine whether these findings reflect methodological errors or actual differences in risk across subgroups.

Geographic representation was also highly limited to Westernized countries. Nearly all studies are conducted in the United States, with only two from Europe, leaving a gap in knowledge regarding low-and middle-income countries where structural drivers of homelessness differ. International studies are needed to clarify how cultural, policy, and geographic contexts shape vulnerability to assault.

Finally, the heavy reliance on cross-sectional designs constrains causal inference. Prospective and longitudinal studies, although resource-intensive, clarify pathways of risk and resilience over time. Greater methodological consistency, including standardized definitions of sexual assault and homelessness, would also enhance comparability.

Addressing these gaps is essential for producing more precise, equitable, and actionable evidence to guide interventions and policies aimed at preventing sexual violence against homeless women.

## 5. Conclusions

The pooled prevalence findings indicate that sexual assault remains a significant and persistent risk for homeless women across diverse contexts. This meta-analysis represents one of the first standardized syntheses of prevalence rates within this population, offering an essential update to the existing literature. By quantifying the scope of the issue and emphasizing associated risk factors, these findings underscore the ongoing vulnerability of homeless women to sexual violence across varying demographic and geographic settings.

Subgroup analyses demonstrated significant heterogeneity (χ² = 152.33, df = 6, p < 0.0001), suggesting that variability in prevalence is attributable to differences in risk profiles and environmental exposures rather than methodological inconsistencies. Among the subgroups analyzed, women with disabilities had the highest prevalence of sexual assault, while HIV-positive women had the lowest. LGBTQ+ individuals and those with mental health conditions exhibited intermediate prevalence rates, though substantial variability persisted. These findings highlight the compounding effect of intersecting social and health-related disadvantages on sexual assault risk.

Several limitations should be acknowledged. Subgroup analyses were constrained by small sample sizes, which may limit the generalizability of results. Additionally, underreporting remains a likely concern, given the stigma, fear, and systemic barriers often faced by homeless women. This may result in an underestimation of the true prevalence. The predominance of studies conducted in North America further restricts the external validity of the findings to other global contexts.

Future research should incorporate data from a broader range of geographical regions and standardized definitions of both sexual assault and homelessness. Consistency in measurement tools would improve reliability across studies. Improved data collection is essential for understanding the intersectional and structural determinants contributing to elevated sexual assault risk among homeless women. From a public health perspective, interventions should focus on improving access to safe reporting mechanisms, enhancing protective measures within shelters and unsheltered settings, and developing trauma-informed, identity-responsive care models tailored to the unique needs of this population.

## Data Availability

All data produced are available online on Open Science Framework

https://osf.io/ny298/overview

## 6. Ethics Declaration

This study is a systematic review and meta-analysis reporting the secondary analysis of aggregated, non-identifiable prevalence data extracted exclusively from previously published, peer-reviewed literature. Review and/or approval by an ethics committee was not needed for this study because the analysis relies entirely on summary statistics and prevalence proportions derived from publicly disseminated research. The authors ensured that all original data included in this meta-analysis were acquired in compliance with the journal’s ethical and consent requirements for secondary analysis of human data. The authors confirm that the primary data sources were peer-reviewed research articles that must have secured appropriate institutional ethical approval and informed consent from their respective participants at the time of their original publication. Since this study utilized only anonymized and aggregated data from the included studies, the necessity for new informed consent from individual participants was obviated.

## 6. Other Information

## 6.1 Funding and Conflicts of Interest

The authors have no known conflicts of interest in the outcomes of this study, nor any incentive to represent findings in a biased manner. As a meta-analysis, the project did not require direct funding.

## 6.2 Acknowledgments

We would like to thank our research assistants, Macarena Sofia Puig, Trevor Anderson, Jacob Duran, Austin Irungu, Rachael Ho, Daisy Kim, Raelene Musharbash, Tyla Williams, Logan Holbrook, Zaneb Mehmood, Nayab Mehmood, Sofia Perez, Gissel, Amy Alarcon, Alessandra Marino, Destiny-Elizabeth Silva-Castro, Kylie Nguyen, Jesus Ernesto Salcedo, Rianna Punzalan, Kelly Tang, Roxanne Carlos Mendoza, Raquel Juliette Tamayo, Ashley Zhao, Anthony Gorman, Andrew, Jocelyn Garcia, Ethan, Natalia Moonesinghe, Simran Athwal, Abigail Restivo, Sarah D. Johnson, Elyse Kong, Carly “Crow” T. Nguyen, Maya Barbosa, Katherine Mejia, Jenny Arrellin, Susannah Jakkula, Mohamed Ibrahim, Oishee Maharatna, Melissa Hernandez-Monroy, Ileana Rodriguez, Melanie Paulette Magallanes, Rhea S., Nipun T, Koyinsola Oyefeso, Aynaya Heysley, Cesar Correra, Ava, Brenda Derin, Nicole, Isaak Lugo, Yacoob, Alondra R, for their contributions to data extraction and screening or management. We are grateful to Qianhan Zhang for statistical analytic recommendations. We also acknowledge our institution, the Valliant Foundation 501 (c), for its support in this research.

## 6.3 Declaration of generative AI and AI-assisted technologies in the manuscript preparation process

During the preparation of this work, the authors used NotebookLM to improve grammatical, spelling, and organizational clarity. After using this service, the authors reviewed and edited the content as needed and took full responsibility for the published article.

## Notes

### Competing Interest Statement

The authors have declared no competing interest.

### Clinical Protocols

https://osf.io/jb57a/overview

### Funding Statement

This study did not receive any funding

## References

1. Basile KC, Smith SG. Sexual violence victimization of women: prevalence, characteristics, and the role of public health and prevention. Am J Lifestyle Med. 2011;5(5):407–17. doi:10.1177/1559827611409512

2. Breiding MJ. Prevalence and Characteristics of Sexual Violence, Stalking, and Intimate Partner Violence Victimization—National Intimate Partner and Sexual Violence Survey, United States, 2011. Morbidity and mortality weekly report Surveillance summaries (Washington, DC : 2002) [Internet]. 2014 Sep 5;63(8):1. Available from: https://pmc.ncbi.nlm.nih.gov/articles/PMC4692457

3. Santa Maria D, Freeden K, Drake S, Narendorf S, Barman-Adhikari A, Petering R, Hsu H-T, Shelton J, Ferguson K, Bender K. Gaps in Sexual Assault Health Care among homeless young adults [Internet]. Am J Prev Med. 2019 Dec 16;58 (2): 191–198. Available from: https://pmc.ncbi.nlm.nih.gov/articles/PMC11006393/

4. Goodman LA, Dutton MA, Harris M. Episodically homeless women with serious mental illness: prevalence of physical and sexual assault. American Journal of Orthopsychiatry. 1995 Oct;65(4):468–78. doi:10.1037/h0079669. Available from : Episodically homeless women with serious mental illness: prevalence of physical and sexual assault - PubMed

5. Yung CR. How to lie with rape statistics: America’s hidden rape crisis. Iowa L. Rev. 2013;99:1197.

6. Gelberg L, Andersen R, Longshore D, Leake B, Nyamathi A, Teruya C, et al. Hospitalizations Among Homeless Women: Are There Ethnic and Drug Abuse Disparities? The Journal of Behavioral Health Services & Research. 2008 Oct 16;36(2):212–32.

7. Harris T, Kintzle S, Wenzel S, Castro CA. Expanding the Understanding of Risk Behavior Associated With Homelessness Among Veterans. Military Medicine. 2017 Sep;182(9):e1900–7.

8. Alessi EJ, Greenfield B, Manning D, Dank M. Victimization and Resilience among Sexual and Gender Minority Homeless Youth Engaging in Survival Sex. Journal of Interpersonal Violence. 2020 Jan 10;36(23-24):088626051989843.

9. Holt VL. Civil Protection Orders and Risk of Subsequent Police-Reported Violence. JAMA. 2002 Aug 7;288(5):589.

10. Warf C, Clark L, Desai M, Calvo R, Agahi G, Hoffman J. Coming of Age on the Streets: Survival Sex Among Homeless Adolescent Females in Hollywood. Journal of Adolescent Health. 2010 Feb; 46(2): 180–2. doi: 10.1016/j.jadohealth.2009.11.088

11. Heslin KC, Robinson P. Langham, Baker RS, Gelberg Lillian. Community Characteristics and Violence Against Homeless Women in Los Angeles County. Journal of Health Care for the Poor and Underserved. 2007;18(1):203–18.

12. Roy AL. Intersectional Ecologies: Positioning Intersectionality in Settings-Level Research. New Directions for Child and Adolescent Development. 2018 Jul 3;2018(161):57–74.

13. Goodman-Williams R, Dworkin E, Hetfield M. Why do rape victimization rates vary across studies? A meta-analysis examining moderating variables. Aggression and Violent Behavior. 2023 Apr 1;71:101839–9.

14. Gangamma R, Slesnick N, Toviessi P, Serovich J. Comparison of HIV Risks among Gay, Lesbian, Bisexual, and Heterosexual Homeless Youth. J Youth Adolesc; 2008.37(4):456–464

15. Ledingham, E. Sexual violence against women with disabilities [Internet]. Am J Prev Med. 2022 [cited 2025 Sep 9]. Available from: https://www.ajpmonline.org/article/S0749-3797(22)00049-6/fulltext

16. Mailhot Amborski A, Bussières EL, Vaillancourt-Morel MP, Joyal CC. Sexual Violence Against Persons With Disabilities: A Meta-Analysis. Trauma, Violence, & Abuse. 2021 Mar 4;23(4):152483802199597.

17. Miles L, Valentine JL, Mabey L, Downing NR. Mental Illness as a Vulnerability for Sexual Assault. Journal of Forensic Nursing. 2022 Jan 18; Publish Ahead of Print.

18. Basile KC, Smith SG. Sexual violence victimization of women: prevalence, characteristics, and the role of public health and prevention. Am J Lifestyle Med. 2011;5(5):407–17. doi:10.1177/1559827611409512.

19. Yung CR. How to lie with rape statistics: America’s hidden rape crisis. Iowa Law Review [Internet]. 2014 Mar [cited 2025 Sep 17];99(3):1197–256. Available from: https://www.researchgate.net/publication/285966206_How_to_lie_with_rape_statistics_ America

20. Loya R.M., Rape as an Economic Crime: The Impact of Violence on Survivor’s Employment and Economic Well-Being. [Internet]. 2014; https://journals.sagepub.com/doi/abs/10.1177/0886260514554291

21. Misganaw AC, Worku YA. Assessment of sexual violence among street females in Bahir-Dar town, North West Ethiopia: a mixed method study. BMC Public Health. 2013 Sep 10;13(1).

22. Calvo F, Watts B, Panadero S, Giralt C, Rived-Ocaña M, Carbonell X. The Prevalence and Nature of Violence Against Women Experiencing Homelessness: A Quantitative Study. Violence Against Women. 2021 Jul 2;28(6-7):107780122110227.

23. Riley ED, Cohen J, Knight KR, Decker A, Marson K, Shumway M. Recent Violence in a Community-Based Sample of Homeless and Unstably Housed Women With High Levels of Psychiatric Comorbidity. American Journal of Public Health. 2014 Sep;104(9):1657–63.

24. Riley ED, Vittinghoff E, Kagawa RMC, Raven MC, Eagen KV, Cohee A, et al. Violence and Emergency Department Use among Community-Recruited Women Who Experience Homelessness and Housing Instability. Journal of Urban Health. 2020 Jan 6;97(1):78–87.

25. Whitbeck LB, Armenta BE, Gentzler KC. Homelessness-Related Traumatic Events and PTSD Among Women Experiencing Episodes of Homelessness in Three U.S. Cities. Journal of Traumatic Stress. 2015 Jul 15;28(4):355–60.

26. Henny KD, Kidder DP, Stall R, Wolitski RJ. Physical and Sexual Abuse among Homeless and Unstably Housed Adults Living with HIV: Prevalence and Associated Risks. AIDS and Behavior. 2007 Jun 19;11(6):842–53.u

27. Riley ED, Shumway M, Knight KR, Guzman D, Cohen J, Weiser SD. Risk factors for stimulant use among homeless and unstably housed adult women. Drug and Alcohol Dependence [Internet]. 2015 Aug 1;153:173–9. Available from: https://www.sciencedirect.com/science/article/abs/pii/S0376871615002598

28. Klarare A, Mattsson E, Gaber SN, Rapaport P, Rosenblad A. Associations Between Violence and Psychological Distress Among Women Experiencing Homelessness: A Cross-Sectional Study. Violence Against Women. 2025 Jun 24;

29. Lucas CL, Harris T, Stevelink SAM, McNamara KA, Rafferty L, Kwan J, et al. Homelessness among Veterans: Posttraumatic Stress Disorder, Depression, Physical Health, and the Cumulative Trauma of Military Sexual Assault. Journal of the Society for Social Work and Research. 2021 Feb 8;12(4). https://www.journals.uchicago.edu/doi/10.1086/712991

30. Wong LH, Shumway M, Flentje A, Riley ED. Multiple Types of Childhood and Adult Violence Among Homeless and Unstably Housed Women in San Francisco. Violence and Victims. 2016;31(6):1171–82.

31. Rosa A da S, Brêtas ACP. A violência na vida de mulheres em situação de rua na cidade de São Paulo, Brasil. Interface - Comunicação, Saúde, Educação [Internet]. 2015 Jun;19(53):275–85. Available from: https://www.scielosp.org/pdf/icse/2015.v19n53/275-285/pt

32. Greene A, Korchmaros JD, Frank F. Trauma Experience Among Women Who Have Substance Use Disorders and are Homeless or Near Homeless. Community Mental Health Journal. 2023 Jul 18;60.

33. Tinland A, Boyer L, Loubière S, Greacen T, Girard V, Boucekine M, et al. Victimization and posttraumatic stress disorder in homeless women with mental illness are associated with depression, suicide, and quality of life. Neuropsychiatric Disease and Treatment. 2018 Sep;Volume 14:2269–79.

